# Late Combination shows that MEG adds to MRI in classifying MCI versus Controls

**DOI:** 10.1101/2021.05.20.21257522

**Authors:** Delshad Vaghari, Ehsanollah Kabir, Richard N. Henson

## Abstract

Early detection of Alzheimer’s Disease (AD) is essential for developing effective treatments. Neuroimaging techniques like Magnetic Resonance Imaging (MRI) have the potential to detect brain changes before symptoms emerge. Structural MRI can detect atrophy related to AD, but it is possible that functional changes are observed even earlier. We therefore examined the potential of Magnetoencephalography (MEG) to detect differences in functional brain activity in people with Mild Cognitive Impairment (MCI) – a state at risk of early AD. We introduce a framework for multimodal combination to ask whether MEG data from a resting-state provides complementary information beyond structural MRI data in the classification of MCI versus controls. More specifically, we used multi-kernel learning of support vector machines to classify 163 MCI cases versus 144 healthy elderly controls from the BioFIND dataset. When using the covariance of planar gradiometer data in the low Gamma range (30-48Hz), we found that adding a MEG kernel improved classification accuracy above kernels that captured several potential confounds (e.g., age, education, time-of-day, head motion). However accuracy using MEG alone (67%) was worse than MRI alone (72%). When simply concatenating (normalized) features from MEG and MRI into one kernel (early combination), there was no advantage of combining MEG with MRI versus MRI alone. When combining kernels of modality-specific features (intermediate combination), there was an improvement in multimodal classification to 75%. The biggest multimodal improvement however occurred when we combined kernels from the predictions of modality-specific classifiers (late combination), which achieved 78% accuracy (a reliable improvement in terms of permutation testing). We also explored other MEG features, such as the variance versus covariance of magnetometer versus planar gradiometer data within each of 6 frequency bands (delta, theta, alpha, beta, low gamma or high gamma), and found that they generally provided complementary information for classification above MRI, provided the frequency band was beta or higher. We conclude that high frequency information in MEG can improve on MRI-based classification of mild cognitive impairment.

## Introduction

Alzheimer’s disease (AD) is an age-related neurodegenerative disorder and a major challenge for healthcare and social care due to its high prevalence and costs. Early detection of AD is critival for treatment and prevenetion, and this requires a robust biomarker that can identify the disease in prodromal stages such as Mild Cognitive Impairment (MCI) (Petersen, 2009). Such biomarkers may also provide disease progression monitoring in clinical trials. Neuroimaging techniques offer a range of potential biomarkers of structural, metabolic and functional changes in the brain related to AD and MCI (Cabeza et al., 2018; Tartaglia et al., 2011; Woo et al., 2017).

Magnetic Resonance Imaging (MRI) is a key such technique, which can be tuned to various tissue properties. The most common of these is the T1-weighted contrast between gray-matter and white-matter – so called “structural MRI” (sMRI) – which can be used to estimate reduction in the grey-matter volume of various brain regions owing to the atrophy in AD. This is a standard approach in assessment of dementia (Frisoni et al., 2010; Nestor et al., 2004). However, AD affects neuronal physiology before cell death and atrophy (Dubois et al., 2016; Han et al., 2012; Jack et al., 2017, 2013). Although functional MRI (fMRI) can be used to measure changes in neural activity and/or connectivity (Agosta et al., 2012; Suckling et al., 2015; van den Heuvel and Hulshoff Pol, 2010; Wang et al., 2006), fMRI only provides an indirect measure of neural function. This is because it relies on the haemodynamic response to neural activity, and is therefore confounded by changes in the brain’s vasculature that occur with age and neurodegenerative disease (Tsvetanov et al., 2019). Furthermore, the slow haemodynamic response limits fMRI to a temporal resolution of seconds.

More direct measures of neural activity can be obtained by Electroencephalography (EEG) and Magnetoencephalography (MEG), which measure the electromagnetic fields produced by dendritic dipoles within active neurons (Hari, MD, PhD and Puce, PhD, 2017; Stam, 2010). These can be sampled at a resolution of milliseconds, revealing a rich repertoire of neural dynamics, including oscillatory rhythms that occur at frequencies between 2 and 100 Hz, such as “alpha” (8-12Hz) and “gamma” (30+Hz), some of which have also been implicated in AD (López-Sanz et al., 2018). MEG offers an advantage over EEG in that the magnetic fields are less distorted and smoothed by the brain-skull interface than are electric fields, resulting in higher spatial resolution (Maestú et al., 2019). We therefore focus on MEG measures of neural activity (during rest) to see if they provide information for MCI classification that is complementary to the structural information in sMRI.

The general advantage of multi-modal integration (combining information from more than one neuroimaging technique) has been appreciated for many years, on the assumption that each modality reveals information about somewhat different aspects of the underlying neural circuity (Engemann et al., 2020; Henson et al., 2011; Kumral et al., 2020; Nentwich et al., 2020) and consistent with findings that combining multiple modalities can improve AD classification (Patel et al., 2008; Polikar et al., 2010). For example, using various different machine learning techniques, Patel *et al*. (2008) and Polikar et al. (2010) all found that combining EEG and sMRI improved diagnosis of AD versus controls compared to using individual modalities alone (though see (Farina et al., 2020)). Note however that the EEG data were collected during an auditory oddball paradigm, rather than the more common resting-state as used here. (Colloby et al., 2016) is the only study we could find that compared resting-state EEG data and structural MRI, but they focused on distinguishing relatively late cases of AD versus Lewy-body dementia, rather than distinguishing relatively early and probable cases of AD (i.e, MCI) versus healthy controls, as done here.

Neuroimaging produces many measurable properties (or “features”) from each participant, such as ∼100,000 voxels in an sMRI image or ∼1,000,000 timepoints in ∼300 sensors in MEG, which normally exceeds the number of participants (typically ∼100). Identifying which features are important for classifying AD therefore benefits from machine learning techniques (Wolfers et al., 2015), such as kernel-based approaches. A kernel is a square matrix containing a measure of the similarity between every pair of participants in their feature values. Classification based on kernels rather than raw data is robust and efficient for high-dimensional pattern classification (Schölkopf and Smola, 2018; Shawe-Taylor and Cristianini, 2004). More specifically, we used Multiple Kernel Learning (MKL) (Gönen and Alpaydın, 2011), which optimises the weighting of kernels from each modality, which is generally better than simply concatenating features across modalities, particularly when the modalities differ in the number of features and/or those features are incommensurate (such as volume in mm^3^ for sMRI versus magnetic field power in fT^2^ for MEG) (Donini et al., 2016; Hughes et al., 2019; Korolev et al., 2016; Liu et al., 2018; Peng et al., 2019; Wee et al., 2012; Youssofzadeh et al., 2017; Zhang et al., 2011). Adding kernels to the classification model is also a better way to accommodate potentially confounding variables (such as the age of MCI and Control cases) than is the more common approach of first adjusting the data (features) for those variables (Dinga et al., 2020; Snoek et al., 2019).

Most importantly, we compared results from combining modalities at three different stages: early, intermediate and late (Figure 1). By early combination, we refer to simple concatenation of the features of each modality, after normalising them by their standard deviation across participants (i.e., to unit-less quantities with comparable numerical range). By intermediate combination, we refer to the typical MKL approach of optimising the weighting of kernels derived from the features of each modality (or confound). By late combination, we refer to the application of MKL to kernels derived from the class predictions after classifiers are run on each modality separately. The latter is closer to the “ensemble learning” philosophy (Kuncheva, 2014) and “stacking” approach used by Engemann et al. (2020).

**Figure 1.**
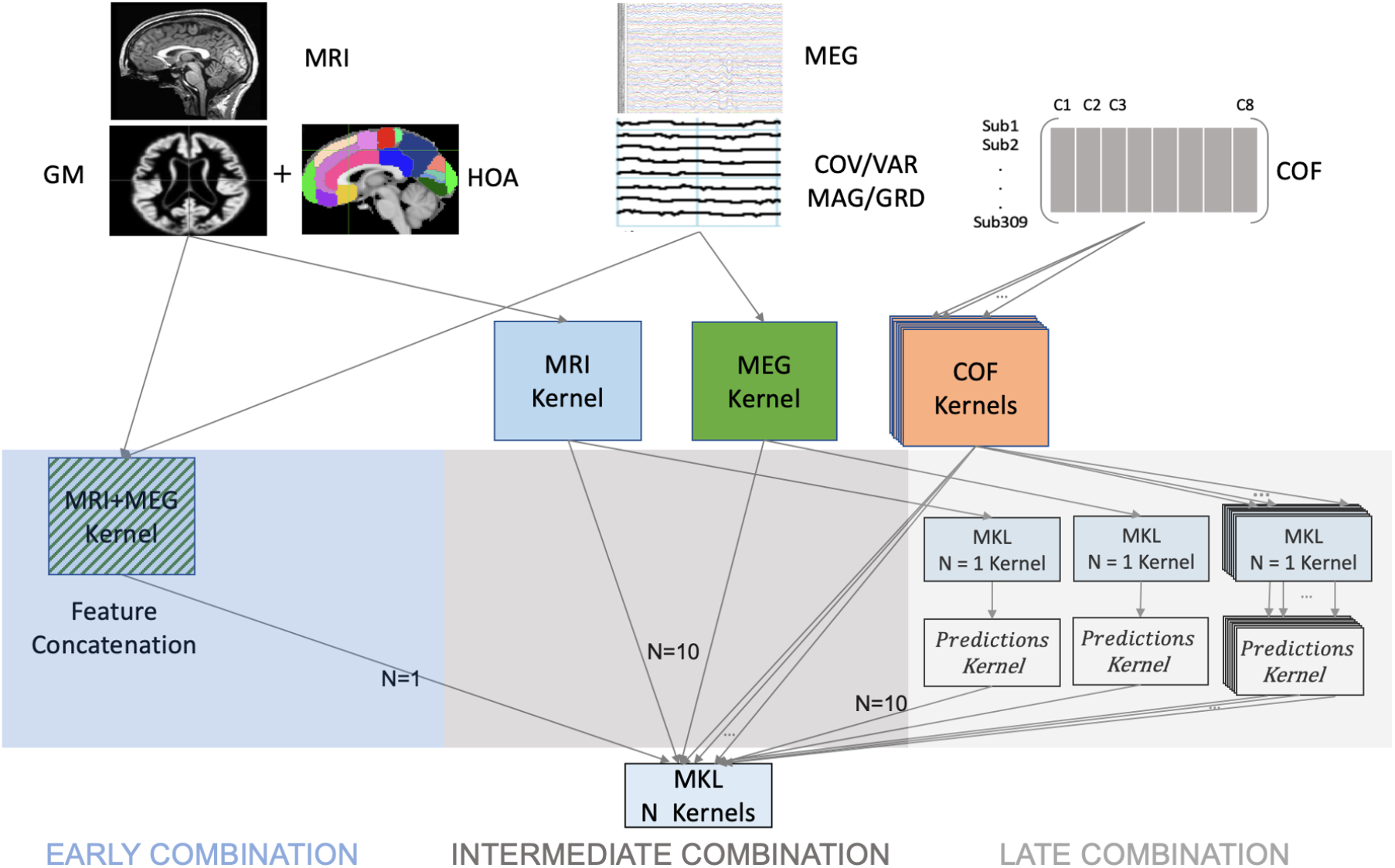
Schematic showing the three different combination stages used here: early (feature concatenation), intermediate (kernel combination) and late (decision combination). N is number of kernels. GM = Gray-Matter, HOA = Harvard-Oxford Atlas, COFs = (potential) Confounds.

## Materials and Methods

### Participants

We included resting-state MEG and T1-weighted structural MRI data from the BioFIND dataset (Vaghari et al., 2021), which includes patients with Mild Cognitive Impairment (MCI) and Healthy Elder Controls (HEC) from two sites: the MRC Cognition & Brain Sciences Unit (CBU) in Cambridge, England, and the Centre for Biomedical Technology (CTB) in Madrid, Spain. The MCI diagnosis was determined with intermediate probability according to the National Institute on Aging–Alzheimer Association criteria (Albert et al., 2011), i.e, given by a clinician based on clinical and cognitive tests, self- and informant-report, and in the absence of full dementia or obvious other causes (e.g, psychiatric). For technical details regarding the MEG and MRI data acquisition, see (Vaghari et al., 2021). After excluding 15 cases without an MRI, and 2 with MRIs with dental artifacts, there were 163 HEC and 144 MCI datasets. A summary of sample characteristics is reported in Table 1, which includes variables that could affect MCI status (such as education) or could affect brain activity in general (such as time of day of testing) or could affect the MEG data specifically (such as head motion or distance from sensors). A small number of missing values were imputed using the mean from non-missing values for each variable. The imputed data are available in the tab-separated value file “participants-imputed.tsv” on the GitHub repository (https://github.com/delshadv/MRI_MEG_Combination).

**Table 1.**
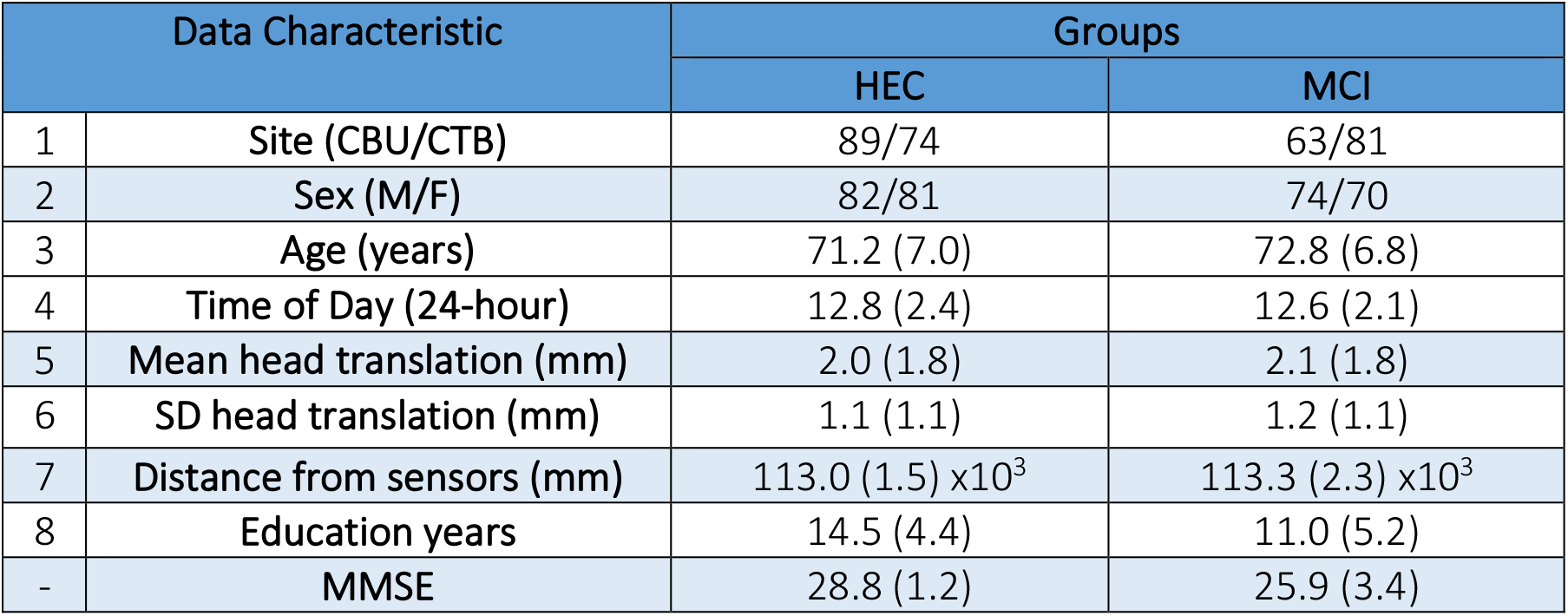
Characteristics of BioFIND participants with clean MRIs. CBU = Cognition & Brain Sciences Unit (Cambridge); CTB = Centre for Biomedical Technology (Madrid). Translation calculated every second from Maxfilter stage (see (Vaghari et al., 2021)). Distance from sensors was calculated after coregistering the MEG to the MRI, and averaging the Euclidean distance between every sensor and each of 2562 vertices on a cortical mesh. SD = standard deviation. MMSE = Mini-Mental State Examination.

### Confounds

There were more HEC cases from the CBU site and more MCI cases from the CTB site, but the number of male/females was close to matched across the two groups. A two-sample T-test confirmed that, on average, the MCI group had lower MMSE scores, as expected from their clinical diagnosis, T(305)=9.63, p<0.001. However, they also had fewer years in education, T(305)=6.41, p<0.001, and were slightly older on average, T(305)=2.09, p<0.05, which may confound any group differences in MRI and/or MEG. While none of the other variables in Table 1 differed significantly between the two groups, T’s<1.46, p>0.14, it is possible that combinations of them could predict MCI status above chance. We therefore included all of them as potential confounds (COFs), except MMSE. The reason we did not include MMSE as a confound is because this cognitive measure informs the MCI diagnosis, so would be circular (biased) to use as a predictor.

### MEG preprocessing

The raw data were de-noised using signal space separation (SSS) implemented in MaxFilter 2.2.12 (Elekta Neuromag) to suppress of environmental noise (Taulu and Kajola, 2005). For more details of max-filtered data and parameters applied to MEG data in BioFIND, please see (Vaghari et al., 2021). The max-filtered (and raw) data are available here: https://portal.dementiasplatform.uk/AnalyseData/AnalysisEnvironment.

The max-filtered data were read into MATLAB 2019a (MathWorks, Natick, MA, USA) using the SPM12 toolbox (http://www.fil.ion.ucl.ac.uk/spm/;(Penny et al., 2006). The minimum duration of resting-state data across participants was 120s, so the first 120s of data was used for all participants. The precise pre-processing steps are provided in the *preproc_meg*.*m* script in the GitHub repository (see above link). In brief, the 120s of data was down-sampled to 500 Hz and band-passed filtered from 0.5-98 Hz (via a high-pass filter followed by low-pass filter). The continuous data were then epoched into 2s segments, and bad epochs were marked using the OSL automatic artefact detection (https://ohba-analysis.github.io/osl-docs/). The number of bad epochs (M=4.66 for MCI and M=4.03 for HEC, out of 60 total) did not differ significantly between groups, T(305)=1.59, p=.11. Non-bad epochs were then concatenated again and bandpass filtered within each of six frequency bands. Visual inspection of the resulting data revealed residual some spikes, particularly in the high frequency bands (low gamma and high gamma). The number of outliers did not differ significantly between groups in any frequency band, T’s<1.33, p’s>.19, except Delta, T(305)=2.60, p=.01, and was always fewer than 1% of samples. These were replaced using the “clip” method of MATLAB’s *filloutliers* function.

### MRI preprocessing and feature extraction

The de-faced, T1-weighted scans were processed in SPM’s DARTEL-VBM pipeline (Ashburner and Friston, 2000), as implemented in the *preproc_mri*.*m* script in the GitHub repository. Each MRI was first segmented into grey matter (GM) and white matter (WM) probability maps. These GM and WM images were then warped to an average template for the sample using diffeomorphic warping in the DARTEL toolbox (Ashburner, 2007), and this template transformed to MNI space. These transformation parameters were then applied to each participant’s GM image, modulated so as to preserve local GM volume, and the GM values resampled into MNI space together with a spatial smoothing by a 1mm FWHM isotropic Gaussian kernel to remove interpolation artifacts. Finally, 110 MRI features were used for classification, representing the mean across voxels within the 110 anatomical ROIs of the Harvard-Oxford Atlas (Kennedy et al., 1998; Makris et al., 1999)^1^.

### MEG Feature extraction

We focused on sensor level features, rather than reconstructing the sources of the MEG data. Firstly, it is unclear whether source reconstruction provides additional information for classification with real data. Since sensor data are a linear combination of source data (mixed through a “forward model”, Hari & Puce, 2017), there no degrees of freedom are gained when estimating source amplitudes. Having said this, when classifying on the basis of non-linear functions of the data, such as the (co)variance (power) features used here, this equivalence between sensor and source features is lost (Sabbagh et al., 2019) It is true that an accurate forward model (which accommodates differences in head position and anatomy) helps align features across participants, and simulations show improvements when estimating sources (Sabbagh et al., 2020), though in practice there are always errors in the forward model and noise in the data that prevent perfect alignment. Indeed, there are normally more sources than sensors, rendering the source estimation problem ill-posed, and requiring additional assumptions to regularize the solution. Secondly, and perhaps more importantly, since an sMRI is necessary to construct an accurate head model, for present purposes of comparing MRI and MEG classification, we did not want information from the MRI to contaminate the MEG features.^2^

The MEG data come from one magnetometer (MAG) and two, orthogonal planar gradiometers (GRD) at each of 102 locations above the head (i.e., 306 channels in total). MAGs measure the component of the magnetic field that is perpendicular to the sensor, whereas GRDs estimate the spatial derivative of the magnetic fields in two orthogonal directions in the plane of the sensor (which is roughly parallel to the scalp). By taking the spatial derivative, GRDs are less sensitive to distant sources, such as environmental noise, and so have a higher signal-to-noise ratio for signals close by, i.e., in superficial cortex. By contrast, MAGs are more sensitive to deeper signals in the brain, but also more susceptible to noise. The best way to combine GRD and MAG data is a matter of contention because they have different SI units (Garcés et al., 2017), but by using separate kernels for each, we do not need to combine them directly.

We focused on the second-order moments of MEG data, i.e., the data covariance across sensors (which is incidentally what most source localization methods are based on). The variances are related to the signal power in each sensor, whereas the covariances are related to the cross-spectral power. Note that sensor covariances capture aspects of both brain activity and connectivity (and can only be attributed solely to connectivity between sources after adjusting for field spread, i.e, linear mixing by the forward model, Engemann *et al*., 2020). This meant 102 features for MAG variance and 204 features for GRD variance, both corresponding to the spatial distribution of power across the scalp, with 5,151 covariance features for MAGs and 20,706 covariance features for GRDs. Note however that our preprocessing of the MEG data (during the MaxFilter stage above) includes a dimension reduction to 69 and 66 components for MAG and GRD respectively (see Supplementary Figure 6), so the rank of the covariance matrices is less. This does not matter for our classification results, since when we reduced the dimensionality further, using principal component analysis (PCA) to calculate the number of components needed to explain 95% of the total variance in the MEG features across participants, the pattern of classification results was hardly changed (see Supplementary Figure 6), with slightly worse accuracies overall.

Each of these features was calculated for 6 frequency bands: Delta [2-4 Hz], Theta [4-8 Hz], Alpha [8-12 Hz], Beta [12-30 Hz], low Gamma [30-48 Hz] and high Gamma [52-86 Hz]. Note that we did not relativize power in each frequency band to the total power (across all frequencies), e.g. by normalizing the timeseries before estimating power (Hughes et al., 2019). While such normalization allows for differences in overall signal strength owing to the proximity of the head (brain) to the sensors, the danger of normalizing power in this way is that it could also remove true power differences between MCI and HEC. We therefore used absolute power, but by including the mean distance between the brain and sensors as a confound, were able to make some allowance for different head positions (after squaring, since the magnetic field strength falls off with at least the square of distance).

### Multimodal Classification

We compared three stages of combining MEG and MRI data: 1. Early combination: features from MRI and MEG are normalised and then concatenated and fed to a single classifier; 2. Intermediate combination: features from MRI and MEG are projected to kernels and MKL then forms an optimal kernel from a function of the original kernels that maximises multimodal classification accuracy; 3. Late combination: MRI and MEG features are fed to one or more classifiers whose continuous-valued outputs (a prediction related to the probability of membership of each class or distance to decision boundary) are then combined in an optimal way, again using MKL (Gönen and Alpaydın, 2011; Kuncheva, 2014; Noble, 2004). According to a common taxonomy of classifier ensemble methods, our late combination is a type of “stacked generalization”, since the outputs of the individual classifiers for each modality are treated as inputs to a meta classifier (here EasyMKL) (Engemann et al., 2020; Wolpert, 1992). It is also worth mentioning that in pattern recognition, early combination is categorised as “feature level” combination whereas late combination is classified in “decision level” combination. Using intermediate or late combinations can be a natural way to control confounds (COF) effect, namely by adding one or more kernels for the (see Discussion). A comparison schematic of these approaches is presented in Figure 1.

Kernel methods project the data features into matrices (kernels) that represent the similarity of the feature vectors between every pair of observations (here, participants), and optimise classification performance based on these kernels (thus each kernel in the present case was a 307 x 307 matrix, regardless of the number of features per modality). Kernels are the basis of several types of classifiers such as MKL (Gönen and Alpaydın, 2011). A linear MKL learns a coefficient vector η that weights each kernel *k* to produce an optimal kernel *K* according to:

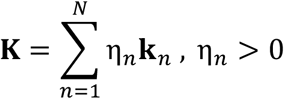

where *n = 1,2,…N* is number of base kernels. Note that we used linear kernels (as well as linear combination), which is recommended when the number of features is larger than the number of participants, i.e., when there is insufficient data to fit more complex nonlinear kernels.

EasyMKL is a MKL algorithm that estimates the relative weighting of each kernel by solving a quadratic optimization problem. Kernels that are not helpful for classification are down-weighted. EasyMKL’s empirical effectiveness has been demonstrated across a large range of kernel numbers (Aiolli and Donini, 2015; Donini et al., 2016). All features were Z-scored across participants before projecting into kernels. We also employed L1 (min-max) normalization of kernels to project all their elements to the interval [0 1]. To ensure that any differences between combination methods did not reflect details of the classifier, the same EasyMKL algorithm was used in all cases (even when only *N=1* kernel in the case of early combination).

The EasyMKL algorithm use a regularization parameter, lambda (λ). A value of λ close to 1 penalizes less informative data, though potentially under-fits data, while a value of λ close to zero does not penalize less informative data, so potentially over-fits data (in that results may be sensitive to outliers in the training data;(Hastie et al., 2009). Here, we use λ=0.1 for Early and Intermediate combination. For Late combination, the same λ=0.1 was used for the first stage of feature kernel combination, but for the second stage of decision combination, we used λ=1 because previous work showed better performance with higher regularization of the second stage relative to first stage (Reid and Grudic, 2009). These values were set a priori (i.e., we did not optimize them using nested-cross-validation). Note however that the better classification for Late combination is not simply because of this larger regularization parameter (in the second stage), because accuracy for Early and Intermediate combination was actually worse, rather than better, when we did a control analysis with λ=1 for Early and Intermediate combination.

Classification performance was estimated using 5-fold cross-validation. Note this applied to both stages of late combination, i.e., performance was always assessed using the untrained fold. Though the overall sample was unbalanced (with more HEC cases than MCI cases), the training set was always selected to be balanced, and the excess HEC participants were assigned to the test set. Given this imbalance in the test set, we report “balanced” classification accuracy, i.e., the mean accuracy across each class separately. Noise simulations in the Supplementary Figure 1 confirmed that our estimation procedure was unbiased.

Cross-validation was repeated 1000 times with random selections of the data, in order to estimate classification reliability. Note that the specific random assignment of participants to training/test sets was matched when comparing different combination methods, meaning that classification accuracies could be directly subtracted for each comparison of interest, such that we could determine the percentage of the distribution of differences in classification accuracies that was greater than zero. This provides an approximation of the reliability of any improvement offered by one approach versus another (e.g., combined MEG and MRI versus MRI alone, or intermediate vs late combination of MEG and MRI).

## Results

### Confounds and single MRI and MEG modalities

Panel a1 in Figure 2 shows the distribution across 1000 permutations of classification accuracies based on 8 kernels, each representing one of the potentially-confounding variables (COFs). The mean accuracy was 58.7%, and above chance (50%) on 98% of occasions. These results come from late combination of the 8 kernels; results using intermediate and early combination are shown in Supplementary Figure 3 and 4. This relatively low level of performance suggests that the two groups were reasonably well-matched in these variables, but the reliable above-chance classification (which may be accidental or causal) provides a baseline to compare with classification using the MEG and MRI features.

**Figure 2.**
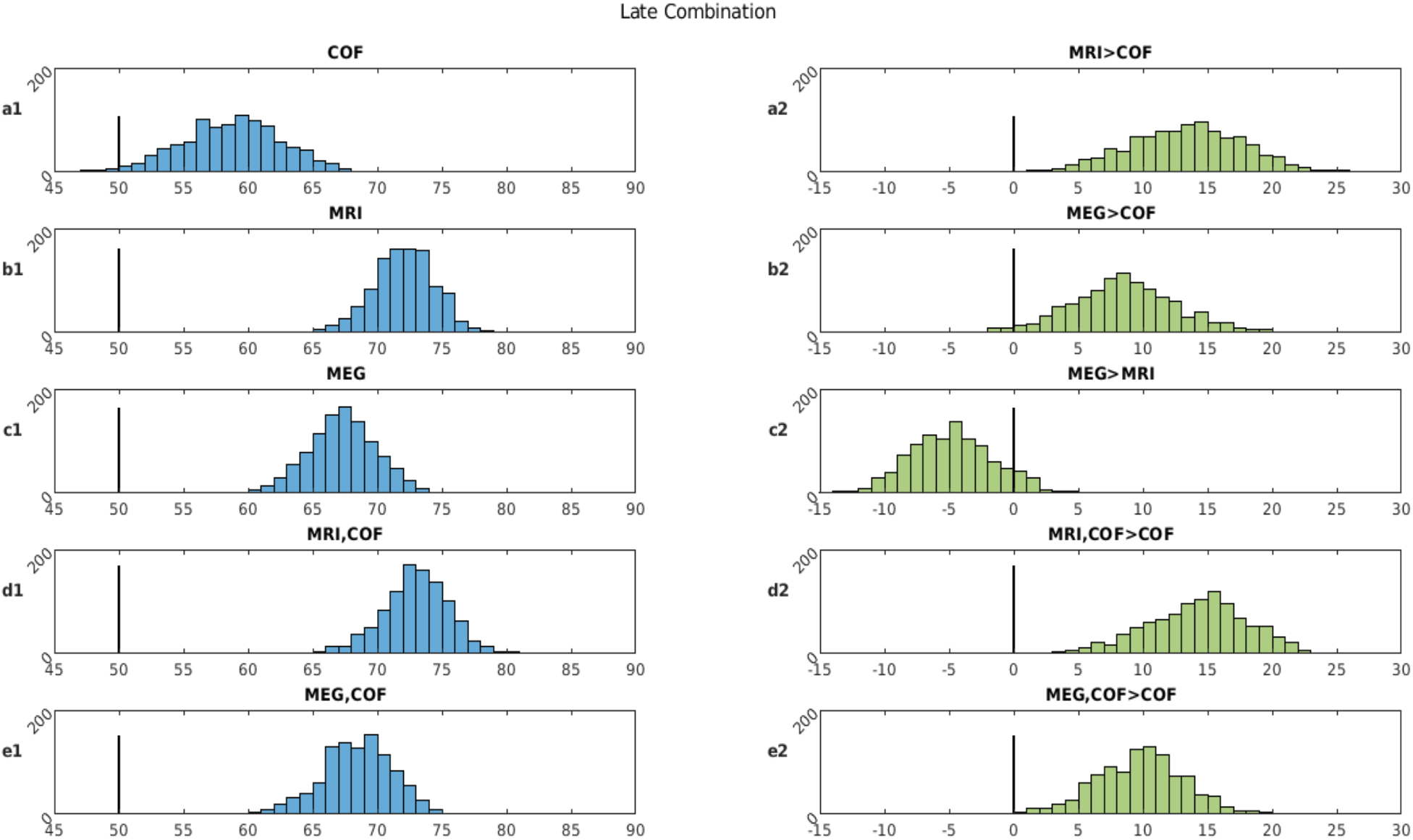
Left column: Classification accuracies (chance = 50%) from 1000 random permutations using late combinations of MRI, MEG (covariance of gradiometers in low-gamma band) and the 8 potential confounding variables. Right column: Differences in classification performance for each permutation when comparing various combinations of features in left column (where 0 = means no difference). “A,B” means combining two (or nine -in presence of confounds) predictions derived from models trained using modality-type A and modality-type B.

**Figure 3.**
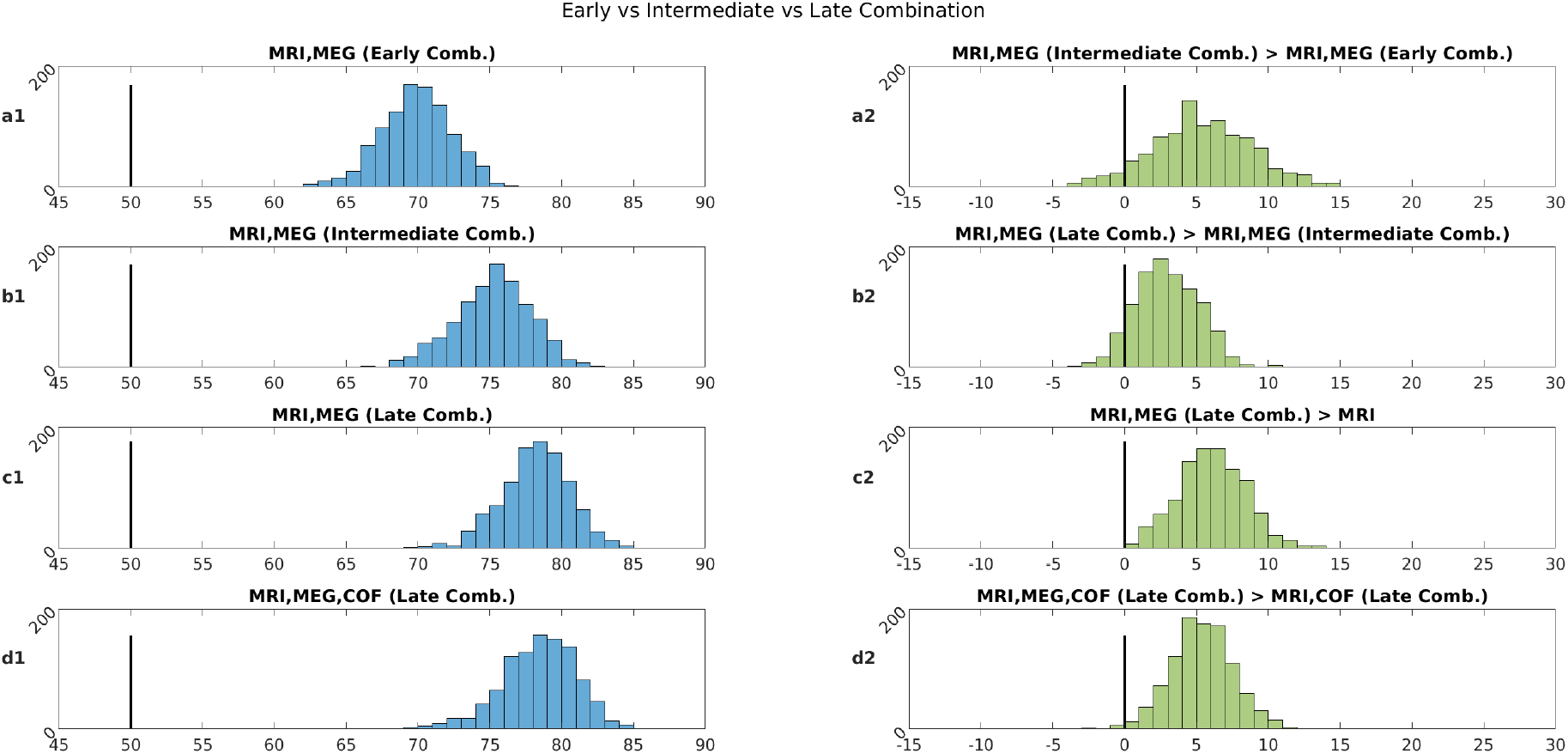
Left column: Classification accuracies (chance = 50%) from 1000 random permutations using MEG, MRI with early, intermediate and late combinations (see methods). Right column: Differences in classification performance for each permutation when comparing various combinations approach (where 0 = means no difference).

For a single kernel based on the MRI features (of GM volume within 110 anatomical ROIs), Panel b1 of Figure 2 shows a mean accuracy of 72.0% (note that there is no distinction between early, intermediate and late combination for a single kernel). To test whether this is a reliable improvement relative to COFs alone, Panel a2 of Figure 2 shows the distribution of differences between accuracies based on MRI versus COFs (when using the same permutations), which showed that MRI was more accurate on 100% of occasions. This demonstrates that MRI provides more information about MCI status than the potential other confounds considered.

For the MEG features, we start with using the covariance across GRDs in the low Gamma range (see later for results using other MEG features). Panel c1 of Figure 2 shows a mean accuracy of 67.3%. While this MEG performance was higher than the baseline provided by the COFs on 98% of occasions (Panel b2), it was lower than for MRI on 91% of occasions (Panel c2). This finding that MRI is generally better than MEG is not surprising, since an MRI is also often used to define the MCI label, i.e., is likely to be biased (see Discussion). The more interesting comparison is whether combining MEG and MRI improves classification relative to MRI alone, which we return to after the next section.

### Adjusting for confounds

Panels d1 and e1 show results from late combination of the COF kernels and either the MRI or MEG kernel respectively. The addition of the COF kernels slightly improves accuracy for both neuroimaging variables, but the more important result is in Panels d2 and e2, which show that these combinations are better than COFs alone on 100% of occasions for both MRI and MEG. This method of combining predictions from confounds (covariates) with those from features of interest (e.g. MRI) is arguably a better way to adjust for confounds than projecting them out of the features of interest themselves (Dinga et al., 2020; Snoek et al., 2019).

### Advantages of early, intermediate and late combination of MRI and MEG

Figure 3 shows classification accuracies when combining the MRI and MEG features at either early, intermediate or late stages. Panel a1 shows that accuracy for early combination is 69.7% which is less than MRI alone (72.0%). This demonstrates that concatenating features from different modalities is not an efficient way to combine them. For Intermediate combination on the other hand, performance is improved on 94% of occasions (Panel a2), with a mean accuracy of 75.2% (Panel b1). This demonstrates that combining feature kernels is better than concatenating features for these data. Late combination improves performance still further, improving on Intermediate combination on 93% of occasions (Panel b2), with a mean accuracy of 78.2% (Panel c1). This demonstrates that combining decision kernels is better than combining feature kernels for these data.

Perhaps the most important result is shown in Panel c2, which shows that late combination of MEG and MRI improves classification accuracy compared to MRI alone on nearly 100% of occasions (99.7%). This suggests that MEG provides information about MCI status that is complementary to that in MRI. The same improvement occurred on 99.7% of occasions when the 8 COF kernels were also added (Panel d1 and d2), showing that this complementary information is also different from anything captured by the potential confounds. The same improvement was also found when massively reducing the ratio of MEG features relative to MRI features by using PCA (Supplementary Figure 6).

### Combining MRI and other MEG features

Given that we selected the COV of GRD in low Gamma as the MEG features, it is possible that the above results are biased by this selection: i.e., when examining all 24 possibilities of variance/covariance (VAR/COV), gradiometers/magnetometers (GRD/MAG) and the 6 frequency bands, it is possible that one or more of them would show better performance when combined with MRI (than MRI alone) due to chance alone. We therefore repeated the above analyses across all frequency bands using both VAR and COV of both GRD and MAG. The results are shown in Table 2, where the top numbers show the mean and standard deviation of classification performance when using late combination of that MEG feature with MRI, and the bottom numbers show the percentage of permutations in which this combination was better than MRI alone. (See Supplementary Table 1 for raw performances for MEG features alone.)

**Table 2.** Exploring the MEG feature space. The top numbers show mean (and SD in brackets) of classification accuracy when combining the relevant MEG feature with MRI, while the percentage below shows the percentage of permutations in which this accuracy exceeded that of MRI alone (72%), where chance = 50%.

| MEG Feature<br>Frequency band | COV<br>of MAG | VAR<br>of MAG | COV<br>of GRD | VAR<br>of GRD |
| --- | --- | --- | --- | --- |
| Delta<br>2-4 Hz | 72.1 (2.4)<br>> MRI on ~50% | 72.1 (2.5)<br>> MRI on ~50% | 72.1 (2.3)<br>> MRI on ~50% | 72.0 (2.3)<br>> MRI on ~50% |
| Theta<br>4-8 Hz | 72.1 (2.4)<br>> MRI on ~50% | 72.1 (2.5)<br>> MRI on ~50% | 72.1 (2.3)<br>> MRI on ~50% | 72.0 (2.4)<br>> MRI on ~50% |
| Alpha<br>8-12 Hz | 72.2 (2.4)<br>> MRI on ~50% | 72.1 (2.5)<br>> MRI on ~50% | 72.0 (2.3)<br>> MRI on ~50% | 72.0 (2.4)<br>> MRI on ~50% |
| Beta<br>12-30 Hz | 72.5 (2.3)<br>> MRI on ~72% | 72.2 (2.5)<br>> MRI on ~50% | 74.2 (2.4)<br>> MRI on ~94% | 72.4 (2.4)<br>> MRI on ~70% |
| Low-Gamma<br>30-48 Hz | 75.1 (2.5)<br>> MRI on ~99% | 73.1 (2.4)<br>> MRI on ~87% | 78.2 (2.4)<br>> MRI on ~100% | 75.8 (2.4)<br>> MRI on ~99% |
| High-Gamma<br>52-86 Hz | 74.5 (2.5)<br>> MRI on ~98% | 73.5 (2.4)<br>> MRI on ~93% | 76.9 (2.4)<br>> MRI on ~99% | 74.6 (2.4)<br>> MRI on ~95% |

Note that for low frequencies (Delta, Theta and Alpha), accuracy when combining them with MRI did not exceed that for MRI alone (72%) – i.e. was only greater on 50% occasions, as expected by chance. For the Beta band, combined accuracy increased slightly, being better than MRI alone on 94% of occasions for the COV of GRD. For low Gamma, multimodal accuracy was consistently better for all four types of feature, suggesting that the choice of VAR/COV or MAG/GRD does not make a big difference (see Supplementary Figure 5 for more evidence). The same was also true of high Gamma. The finding that multimodal combination improved accuracy for over a third of the feature space suggests that the improvement is not a fluke occurrence.

## Discussion

The main finding of the present study was that certain features of MEG resting-state data, particularly in the low and high Gamma frequency range, improve classification of MCI patients versus healthy controls when combined with features from structural MRI data. While classification accuracy with MEG alone never exceeded that for MRI alone, this is not necessarily surprising, since an MRI is typically used by the clinicians to support the diagnosis of MCI (i.e., giving MRI an unfair advantage). The important result was that combining MEG with MRI improved classification relative to MRI alone. This indicates that MEG contains complementary information about MCI. This information might include changes in functional activity and/or connectivity that precede structural change, at least as measured by regional gray-matter volume as here (Dubois et al., 2016; Han et al., 2012; Jack et al., 2017, 2013).

The second main finding of the present study was that late combination of MEG and MRI data was best for multimodal classification, i.e., better than intermediate or early combination. Late combination here refers to combining the class predictions of classifiers trained on each modality separately, analogous to ensemble learning (Kuncheva, 2014). Permutation tests showed that this combination at the “decision-level” reliably improved accuracy relative to intermediate combination at the “feature-level”, where kernels derived from the features of each modality were combined directly. As expected, intermediate combination via kernels was in turn better than simply concatenating (normalized) features into a single kernel. Note that these findings were obtained when the same classification algorithm (EasyMKL; (Aiolli and Donini, 2015) -i.e., multi-kernel learning of support vector machines -was used for each level of multimodal combination.

A third finding was that, while the choice of using either the variance or covariance of MEG magnetometers or planar gradiometers did not make a big difference to multimodal classification accuracy, it was important to consider high-frequency components of the MEG data, specifically the low (30-48 Hz) or high (52-86 Hz) Gamma range. Accuracy when using the Beta range did not produce such a large improvement over MRI alone, while that for Alpha, Theta or Delta provided no improvement. This suggests that the important information for MCI classification exists in frequencies above approximately 30 Hz (see below for further discussion).

A fourth outcome was the demonstration that potential confounds in any classification problem can be accommodated within a multi-modal combination approach, in which confounds are combined with the features of interest during classification (note this applies whether the combination is early, intermediate or late). To “control for” such confounds, one can show that classification accuracy with the combined data reliably exceeds that for the confounds alone. This is better than the common approach of first adjusting the features of interest by the confounds (e.g., by projecting out of the features of interest anything that can be explained by a linear combination of the confounds, i.e., before creating the feature kernels), because it takes into account the potential shared dependency between the features of interest and the confounds in their ability to predict the class (Dinga et al., 2020).

Furthermore, some classifiers are non-linear, and therefore potentially sensitive to effects of confounds that cannot be removed from the features by linear methods.

### Complementary information in MEG/EEG for MCI classification

Previous studies have shown that EEG and/or MEG provide complementary information beyond structural MRI, whether that be in predicting age (Engemann et al., 2020) or classifying types of dementia (Colloby et al., 2016; Patel et al., 2008; Polikar et al., 2010). Some of these have used evoked EEG responses during tasks (e.g., auditory oddballs (Patel et al., 2008; Polikar et al., 2010), which may provide further information on neurodegeneration than the resting-state data used here, though resting-state data are at least more common and easier to obtain, particularly in patients who might struggle with some tasks. In the data paper describing the format and access to the BioFIND dataset (Vaghari et al., 2021), we reported validation analyses that showed that MEG power across all sensors, or power across all cortical sources, or connectivity between all pairs of sources (based the correlation of the power envelopes) achieved similar classifcation accuracies between 63-67%, comparable to the figure of 67% here for MEG alone (at least for COV of GRD in low Gamma). However, in that paper, we did not compare MEG classification with that from MRI. In a previous paper describing the larger BioFIND project (Hughes *et al*., 2019), we did report preliminary findings that intermediate combination of MEG and sMRI improves classification, using a subset of roughly half (N=168) of the present cases. However, this finding used different features (interpolated 3D scalp-frequency power images for MEG and voxel-level GM images for sMRI) and did not establish the reliability of the improvement using permutation. The present work confirms the reliability of these findings, and extends the approach to demonstrate the added value of late combination of modalities, and the potential value of covariance, rather than power, across MEG sensors within certain frequency bands.

It is important to note that we have only used one type of structural brain information – namely gray-matter volume as estimated within 110 ROIs from a T1-weighted MRI. Other MRI modalities (e.g., T2-weighted MRI, diffusion-weighted MRI, magnetic resonance spectroscopy, MRS) – or indeed even other ROI selections or different pre-processing of the current T1-weighted images – might enable better MCI classification, to the extent that MEG no longer adds further improvement. Furthermore, other imaging techniques like PET might do better still, given their ability to measure neurotransmitters or molecular pathologies directly related to AD. However, our main purpose here was to compare MEG with the most common type of brain image available on MCI patients, and most common type of informal inspection done by clinicians, i.e. looking for gray-matter atrophy.

We cannot tell whether the complementary information provided by MEG here relates to the fact that it measures brain activity/connectivity rather than brain structure, or simply that it is a completely different type of brain measurement with different spatial and temporal properties. Indeed, it would be interesting to apply the present multimodal classification approach to fMRI data, to see if MEG continues to provide more information than fMRI. If MEG does not improve classification beyond fMRI, then the key additional information for better MCI classification might simply be the inclusion of measures of brain function rather than structure; alternatively, if MEG does improve beyond fMRI, it may be that MEG captures neural activity more directly (bypassing the vascular confounds in fMRI) and/or neural activity that is beyond the temporal resolution of fMRI. Unfortunately resting-state fMRI is not available in BioFIND, but is likely to be available together with MEG/EEG and sMRI in future cohorts being studied around the world. Likewise, one could test whether the improved spatial resolution of MEG over EEG also provides additional information for MCI classification, and whether task-based MEG/EEG provides additional information beyond the resting-state data used here.

### Multimodal classification approaches

Our best classification accuracy of 78% when using MEG and MRI may not seem particularly impressive, for example relative to figures reported in other papers using different modalities and datasets. However, it is important to note that our aim was not simply to achieve the best classification possible. For example, while we found similar results after PCA to reduce the feature dimensionality (Supplementary Figure 6), we could have employed more sophisticated feature selection approaches that might have improved classification accuracy, particularly given the large number of MEG features relative to participants, or we could have tried to minimize effects of field spread on our second-order (covariance) features by employing spatial filtering or Riemannian Embeddings (Sabbagh et al., 2020). Furthermore, we could have used neuroscientific knowledge to select features, for example based on knowledge that the medial temporal lobes include some of the structures affected in the earliest stages of AD (Frisoni et al., 2010; Shi et al., 2009). We could have even optimized the EasyMKL hyper-parameter (e.g., using nested cross-validation) instead of choosing a fixed value at the outset. Rather, our aim was only to compare the relative performance of different modalities and different methods of combining those modalities, while holding other factors constant.

It is also important to note that there are likely to be errors in our MCI labels, since they are based on clinician’s diagnoses. For example, some of the participants labelled as MCI in the BioFIND dataset may in fact have healthy brain structure and/or function (i.e., no evidence of early AD), but just perform poorly on cognitive tests because of other reasons like depression. Indeed, an appreciable proportion of clinically diagnosed MCI cases later turn out to have no detectable AD pathology (Petersen, 2009). Conversely, some participants labelled as healthy controls may have had early AD and impairments of brain function and/or structure, but performed normally on cognitive tests because of high pre-morbid ability or some form of “cognitive reserve” (Stern, 2009). This places an upper limit on how well MEG and/or MRI could classify the present data. These issues can only be resolved by longitudinal follow-up, possibly with additional biomarkers (e.g., CSF Tau levels) and ultimately post mortem examination to confirm who had AD. While follow-up data are available on some of the BioFIND participants, and may be added in future, there was not currently enough to be able to utilize in the present analyses.

It is important to note that, while the BioFIND dataset may be the largest sample of MCI patients and controls with MEG data, it is still small for machine-learning approaches, relative to the potential number of features (e.g., 20,706 for covariance of planar gradiometers). This dearth of training data may also explain why higher classification accuracies have been reported for other neuroimaging markers (e.g., sMRI) for which larger databases exist, such as ADNI (http://adni.loni.usc.edu/about/). As a reference, using MRI alone on the ADNI database of N=1409 cases (294 patients with probable AD, 763 patients with MCI, and 352 healthy controls), (Basaia et al., 2019) reported an accuracy of 76% for stable MCI cases, using sophisticated deep neural net classifiers. Interestingly, this is only 4% more than achieved here using standard SVMs on a smaller set of MRI cases, though these authors did achieve a higher performance of 87% for those MCI cases who subsequently converted to confirmed AD, reinforcing the above point of heterogeneity within typical MCI cases.

Furthermore, in situations with many more features than cases, overfitting is likely (Cawley and Talbot, 2007; Cristianini and Shawe-Taylor, 2000; Han and Jiang, 2014). This is a situation where late combination might increase generalisation to new datasets, by virtue of the combination of decisions being more robust to over-fitting (Kuncheva, 2014; Wolpert, 1992). Indeed, the simulations in Panel d of Supplementary Figure 2 confirm that Late combination can be better than Intermediate combination when more noise features are added i.e., is more robust against addition of weak (in terms of accuracy) classifiers trained on noisy features. Note however that Late combination is not always better than Intermediate combination, and multi-kernel combination is not always better than simple feature concatenation (Early combination), as can be seen by comparing accuracy for the 8 COFs in Figure 2 with Supplementary Figures 3 and 4, i.e., in situations with relatively low numbers of features. These results depend on the regularisation parameters used, which would ideally be optimised as a function of the specific features too, but is beyond the present remit.

### Optimal MEG features for MCI classification

Our finding that low and high Gamma frequencies provide the information that is complementary to MRI is consistent with some previous M/EEG studies of MCI or genetic risk that highlight the importance of the gamma band (Bajo et al., 2010; Luppi et al., 2020; Missonnier et al., 2010; van Deursen et al., 2008) (Koelewijn et al., 2019). However, other M/EEG studies of MCI (Garcés et al., 2013; Hughes et al., 2019; López et al., 2014; López-Sanz et al., 2018; Maestú et al., 2019; Nakamura et al., 2018) have argued that the alpha band is best for distinguishing MCI versus controls. One possibility is that the information about MCI status provided by Alpha power is correlated with the gray-matter atrophy provided by MRI, which is why we did not find any improvement when combining MRI with MEG covariance in this frequency band. However, it is worth noting that best classification accuracy with Alpha alone (∼60%) was still considerably lower than for Gamma alone (∼69%; Supplementary Table 5). Another reason for this discrepancy in the literature may reflect the features used, for example, the amplitude or frequency of the prominent Alpha peak in MEG and EEG power spectra is a type of feature that is not simply (e.g. linearly) derivable from the Alpha covariance matrix used here. Other discrepancies may owe to the use of different and relatively small samples (N<100), possibly with different definitions of MCI patient, and to the wide range of methodological approaches (Yang et al., 2019). Thus, we do not wish to claim that Gamma frequencies, or more specifically covariance of planar gradiometers, are always the best MEG features to use for MCI classification. We only used the sensor covariance matrix as a simple but inclusive measure of functional activity and connectivity, and explored the range of frequency bands because of prior evidence that frequency matters. Nonetheless, future studies could use the present framework to ask whether specific MEG features offer additional information about MCI over other MEG features.

## Supporting information

All supplementary Material

## Data Availability

Data are available on DPUK website given in Vaghari et al (2021) MedRXiv paper

https://portal.dementiasplatform.uk/CohortDirectory/Item?fingerPrintID=BioFIND

https://github.com/delshadv/MRI_MEG_Combination

## Supplementary material

Supplementary material is available at xxx online.

## Code Availability

The custom written codes to implement all validation analyses is available on GitHub (https://github.com/delshadv/MRI_MEG_Combination). All MEG and MRI features as well as other derived variables are available in comma separated value (.csv) files in “derived” directory within the repository. The raw data are available on the DPUK website (https://portal.dementiasplatform.uk/Apply) cited in main paper.

## Acknowledgements

We thank all the participants who contributed their time to these studies, and the scientists who helped collect the data. We thank in particular Laura Hughes for help collecting the Cambridge data, James Rowe for clinical advice, Denis Engemann for helpful comments on the paper, and Ricardo Bruña Fernandez and Fernando Maestú for collating the Madrid data. This work was supported by: EU JNPD (MR/P502017/1) and MRC (SUAG/046 G101400).

## Footnotes

1 ROI numbers 111-116 in the HOA are spurious or non-cortical.

2 There are data-driven alternatives, for example to adjust the sensor covariance to more accurately reflect source covariance using Riemannian Embeddings (Sabbagh et al., 2020), which could be tried in future and compared with present results.

