## Supplementary material for "Late Combination shows that MEG adds to MRI in classifying MCI versus Controls": All supplementary Material

### Noise Simulations

We conducted a number of simulations to check the validity of our multi-modal combination algorithm and cross-validation scheme. We used the same MCI/HEC labels as the real data in the main paper, i.e. 307 observations. This is performed by the MATLAB script *noise_sim.m* in the GitHub repository.

Supplementary Figure 1 shows the results from 1000 realisations of simulated data; panels on the left come from Intermediate combination while panels on the right come from Late Combination. First, we simulated 1000 "noise" features (drawn from a zero-mean Gaussian with standard deviation of 1). Panels a1 and a2 show that performance was centred on 50%, as expected. This demonstrates that our cross-validation scheme is unbiased.

Next we simulated a single feature based on the (z-scored) MMSE values, as a reference "signal" feature. Panels b1 and b2 show identical mean performance of 73.6% for both Intermediate and Late combination, as expected when only one kernel.

To demonstrate that Early combination is not optimal, we concatenated the 1000 noise features to the MMSE signal feature, and panels c1 and c2 show performance dropped to 54.5%, reflecting the difficulty in finding the signal among so many features.

However, when combining the signal feature and noise features via separate kernels (i.e., 1001 kernels), Panels d1 and d2 show that performance improved related to Early combination, to 58.2% for intermediate and 70.7% for late combination (though still below the reference levels in Panels b1 and b2 with only the MMSE signal).

Finally, Panels e1 and e2 show the results from 4 "signal" kernels – the original MMSE plus education, age and sex. Performance is now improved to 73.7% for Intermediate combination (Panel e1) and 76.6% for Late combination (Panel e2).

Supplementary Figure 2 shows the distribution of differences between Intermediate and Late combination. As expected, there is no difference when a single kernel in Panels a, b and c, but there is a reliable improvement (on over 90% of occasions) for Late combination when there are 1001 (Panel d) or 4 (Panel e) kernels.

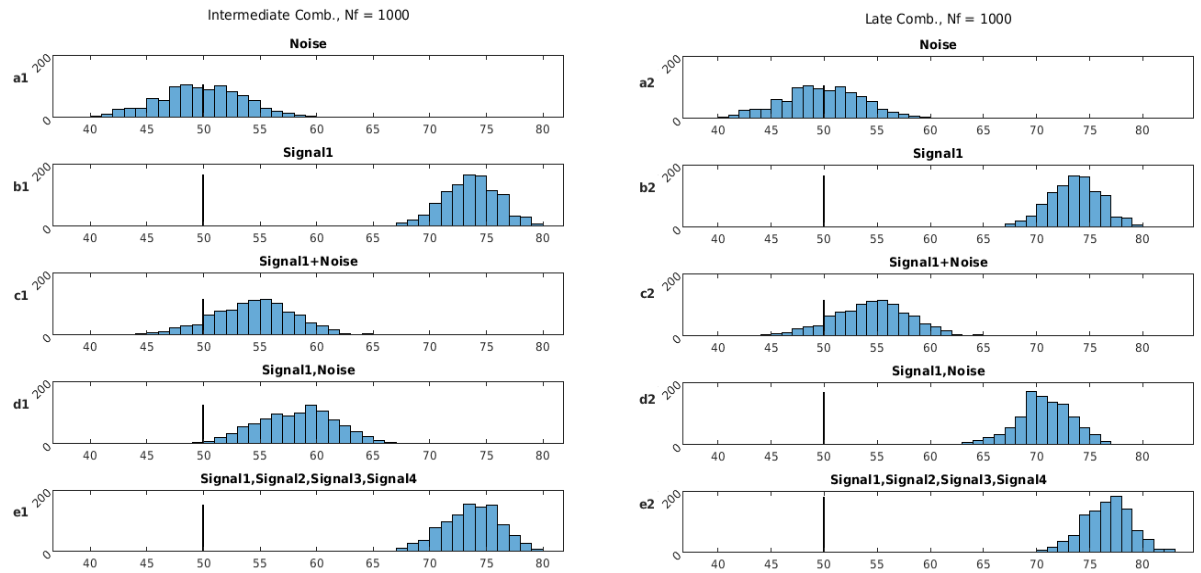


Supplementary Figure 1 Classification accuracies (chance = 50%) from 1000 noise realizations of various kernels (see text) for Intermediate (left column) and Late (right column) combination. "Noise" refers to 1000 random features drawn from a unit normal distribution; "Signal1" refers to MMSE score; "Signals 2-4" refer to Education, Age and Sex. "A,B" means combining either feature kernels (left column) or prediction kernels (right column) derived from models trained using features A and B; "A+B" means concatenation of features from A and B.

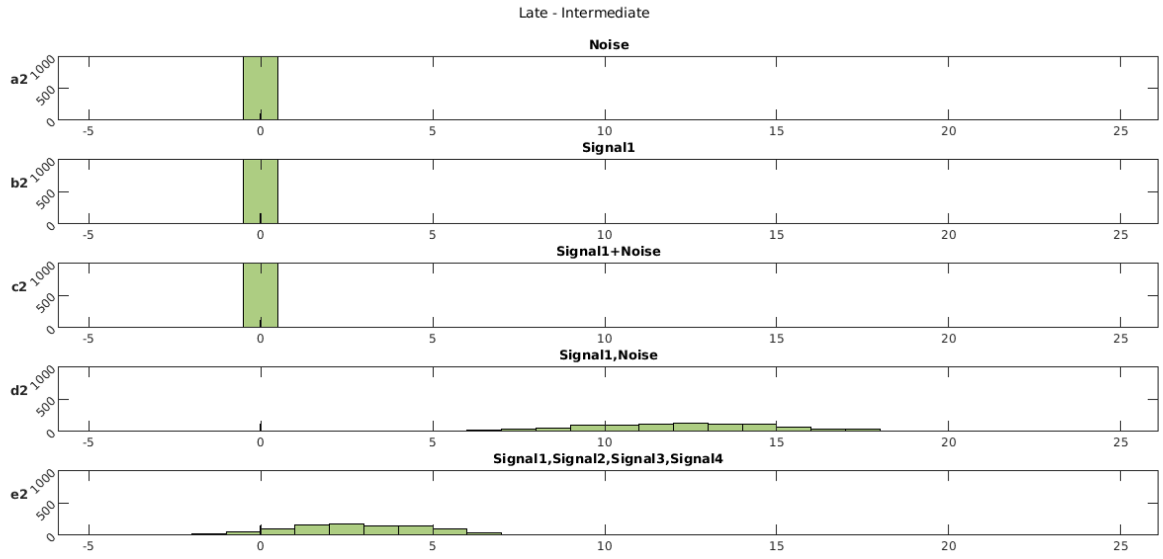


Supplementary Figure 2. Differences in Late versus Intermediate classification accuracies (chance = 0%) from 1000 noise realizations of various kernels (see text). "Noise" refers to 1000 random features drawn from a unit normal distribution; "Signal1" refers to MMSE score; "Signals 2-4" refer to Education, Age and Sex. "A,B" means combining either feature kernels (left column) or prediction kernels (right column) derived from models trained using features A and B.

### Comparison of different modalities using Early or Intermediate combination.

Figure 2 of the main paper shows classification performance for the 8 confounds (COFs), MRI and MEG (covariance of low Gamma in gradiometers) using Late Combination; Supplementary Figures 3 and 4 show the same results for Early and Intermediate combination respectively (note the COFs do not include the MMSE score used Supplementary Figures 1-2, owing to circularity in the MCI classification explained in the main paper).

For Early combination in Panel a1 of Supplementary Figure 3, the concatenation of COFs produced mean performance of 65.8%, which is actually above that for Intermediate (Supplementary Figure 4) and Late (main Figure 2) combination using a separate kernel for each confound. The reason for this is considered below. As a result, though MRI still improves classification above COFs on 97% of occasions (Panel a2), MEG only improves classification on 66% of occasions (Panel b2).

The results for MRI or MEG alone in Panels b1, c1 and c2 are identical to Intermediate and Late combination because only a single kernel is involved. More interesting is the Early combination of MRI and MEG with COFs (Panels d1-e1): while adding MRI to COFs still improves classification on 100% of occasions (Panel d2), adding MEG to COFs only improves on 66% of occasions, unlike for Intermediate (Supplementary Figure 4) and Late (main Figure 2) combination, where MEG improved on 95% and 100% of occasions respectively.

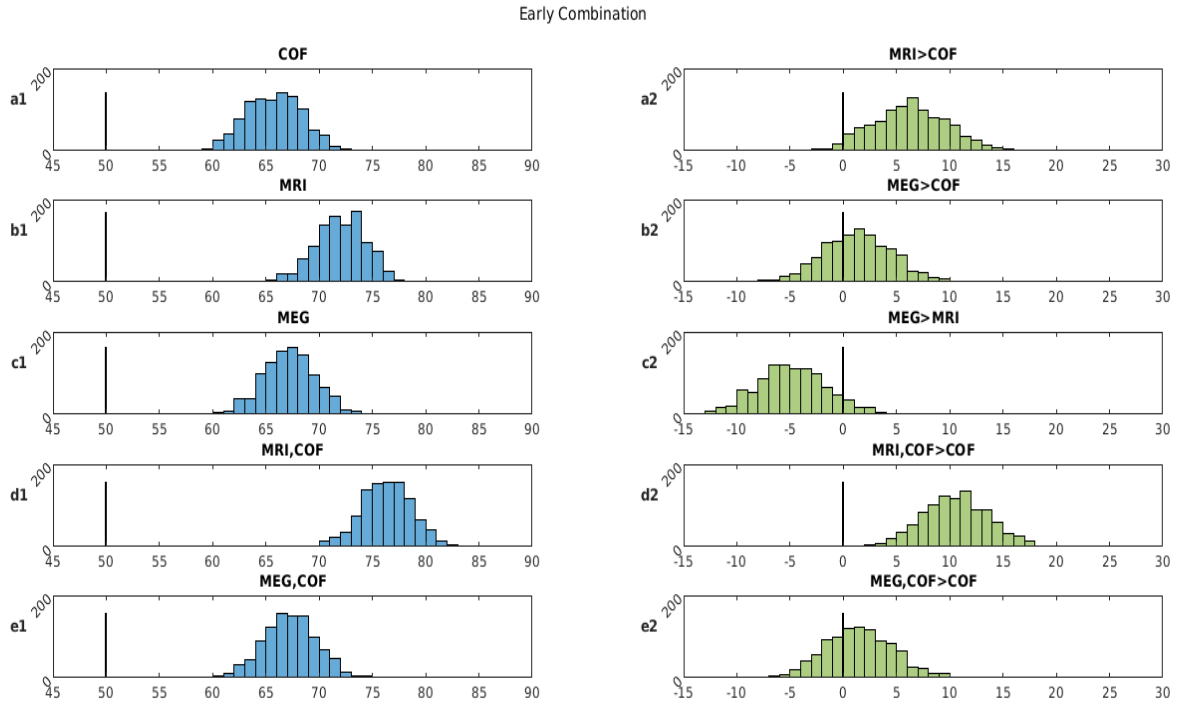


Supplementary Figure 3 Left column: Classification accuracies (chance = 50%) from 1000 random permutations using Early combinations of MRI, MEG (covariance of gradiometers in low-gamma band) and the 8 potential confounding variables. Right column: Differences in classification performance for each permutation when comparing various combinations of features in left column (where 0 = means no difference). “A,B” means combining two (or nine - in presence of confounds) predictions derived from models trained using modality-type A and modality-type B.

Supplementary Figure 4 shows the same set of comparisons as in Supplementary Figure 3, but now for Intermediate rather than Early combination. As noted above, performance actually drops to 64.7% for the 8 COFs (Panel a1) relative to Early combination, though is still above that for Late combination (58.7%) in Figure 2 of the main paper. This relative performance for the COFs kernels of Early > Intermediate > Late is opposite to the case when combining MRI and MEG kernels, as shown in Figure 3 of the main paper, where Late > Intermediate > Early. This demonstrates that Late combination does not always improve over Intermediate combination, and combining kernels (using MKL) does not always improve over simple feature concatenation. The relative advantage of each combination method depends on the number and nature of the features, as well as the regularisation parameters used (which could be adjusted according to the number of features, but which we fixed here to enable comparison).

As above, the combination method has no effect when only one kernel in Panels b1, c1 and c2. More important is, when combining MEG with COFs (Panel e1), performance is improved relative to Early combination, such that adding MEG to COFs (Panel e2) now does reliably improve classification on over 95% of occasions (Panel e2 of Supplementary Figure 3).

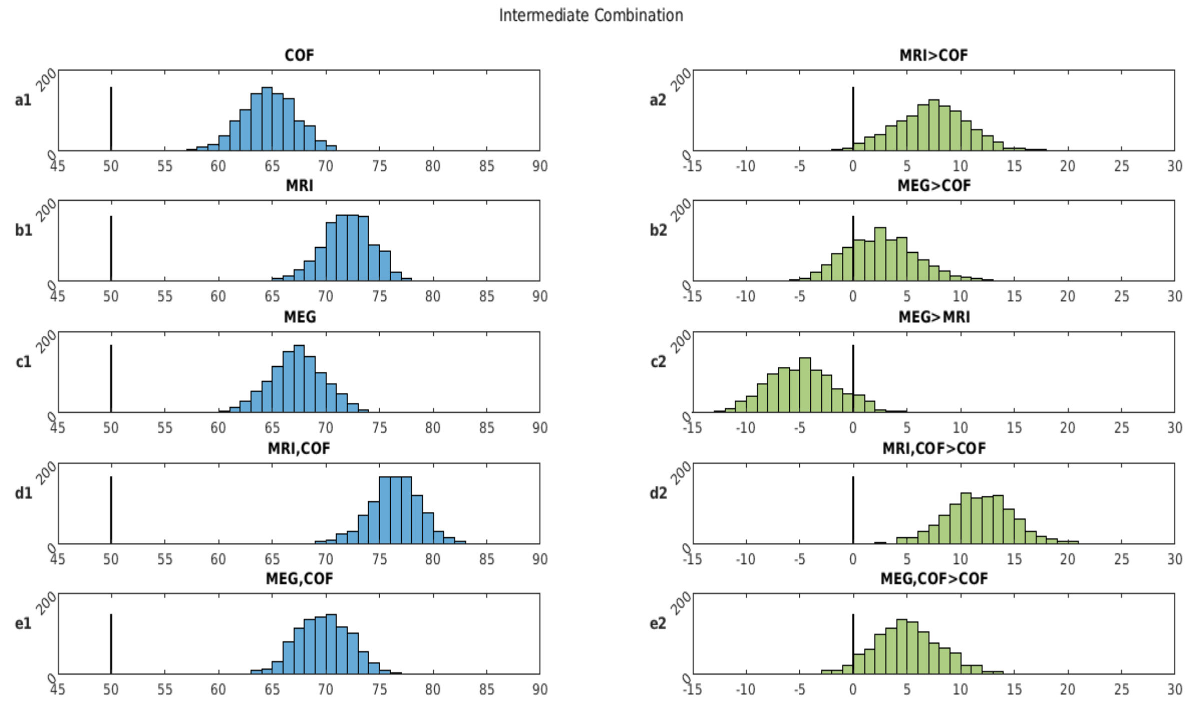


Supplementary Figure 4 Left column: Classification accuracies (chance = 50%) from 1000 random permutations using Intermediate combinations of MRI, MEG (covariance of gradiometers in low-gamma band) and the 8 potential confounding variables. Right column: Differences in classification performance for each permutation when comparing various combinations of features in left column (where 0 = means no difference). "A,B" means combining two (or nine - in presence of confounds) predictions derived from models trained using modality-type A and modality-type B.

### Classification Accuracies for all MEG features

Table 1 in the main paper shows the percentage of occasions when combining MRI with various MEG features improve classification relative to MRI alone; Supplementary Table 1 shows the raw classification accuracies.

| MEG Feature / Frequency band | COV of MAG | VAR of MAG | COV of GRD | VAR of GRD |
|---|---|---|---|---|
| Delta 2-4 Hz | 56.3 (2.6) | 54.7 (2.4) | 60.9 (3.0) | 59.8 (2.6) |
| Theta 4-8 Hz | 60.2 (2.5) | 56.3 (2.7) | 60.5 (2.7) | 58.7 (2.7) |
| Alpha 8-12 Hz | 59.7 (2.6) | 55.3 (2.6) | 60.1 (2.5) | 56.7 (2.5) |
| Beta 12-30 Hz | 64.6 (2.6) | 59.3 (2.8) | 67.6 (2.5) | 65.2 (2.7) |
| Low-Gamma 30-48 Hz | 67.4 (2.5) | 63.8 (2.5) | 67.3 (2.5) | 67.9 (2.5) |
| High-Gamma 52-86 Hz | 68.9 (2.4) | 65.7 (2.6) | 68.1 (2.5) | 66.7 (2.6) |

*Supplementary Table 1. Exploring the MEG feature space. The numbers show mean (and SD in brackets) of classification accuracy from 1000 permutations of 5 fold cross-validation of various MEG features in sensor level.*

Supplementary Figure 5 shows the distribution of classification accuracies across 1000 permutations using Late combination for low gamma band [30-48 Hz], as a function of VAR/COV and MAG/GRD. While there was some evidence that covariance classified better than variance (on 73% of occasions in Panel a2) and that gradiometers classified better than magnetometers (on 86% occasions in Panel c2), there was no convincing evidence that the information was complementary, in that combining COV with VAR did not improve classification reliably above VAR alone (Panel b2) and combining GRD with MAG did not improve classification reliably above MAG alone (<90% of occasions) (Panel d2). This is not necessarily surprising, since VAR and COV both capture a mixture of source activities (in addition to any connectivity also contributing to COV), and that the dependency between MAG and GRD is further increased by the fact that MaxFilter reconstructs them from back projection of the same SSS components (Taulu et al. 2005; Taulu and Kajola 2005).

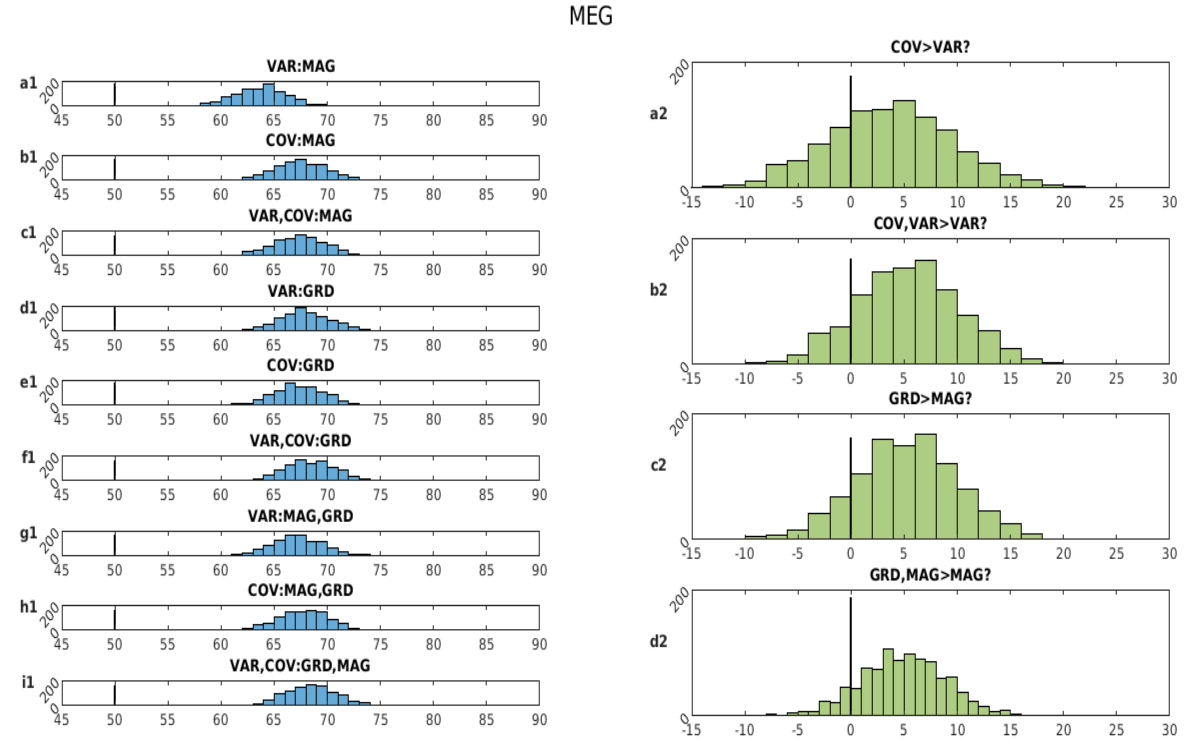


**Supplementary Figure 5 Left column: Classification accuracies (chance = 50%) from 1000 random permutations using Late combinations of different MEG features/sensor. Right column: Differences in classification performance for each permutation when comparing various combinations of features in left column (where 0 = means no difference). "A,B" means combining two predictions derived from models trained using sensor or feature-type A and sensor or feature -type B.**

Supplementary Figure 6 shows classification accuracies when combining the MRI and MEG features at either early, intermediate or late stages, like Figure 3 in main paper, but after applying PCA to reduce dimensions to the number needed to explain at least 95% of the variance in the features across participants. For MEG (covariance of gradiometers in low gamma), this number was 57; for MRI, it was 11.

Panel a1 shows that the mean accuracy for early combination of 69.4% is slightly lower than that without PCA (69.7%). For Intermediate combination, the mean accuracy of 75.6% is slightly better than that without PCA (75.2%) and is improved on 95.6% of occasions (Panel a2). The mean accuracy for late combination with PCA is 76.3%, less than the 78.2% without PCA, and improving on Intermediate combination on 64.1% of occasions (Panel b2). There is also a reduction in performance of MRI alone, from 72.0% without to 71.3% with PCA. Most importantly, with PCA, late combination of MEG and MRI is still better than MRI alone on 100% of occasions (panel c2), and the same results occurred when COFs were added (100% of occasions).

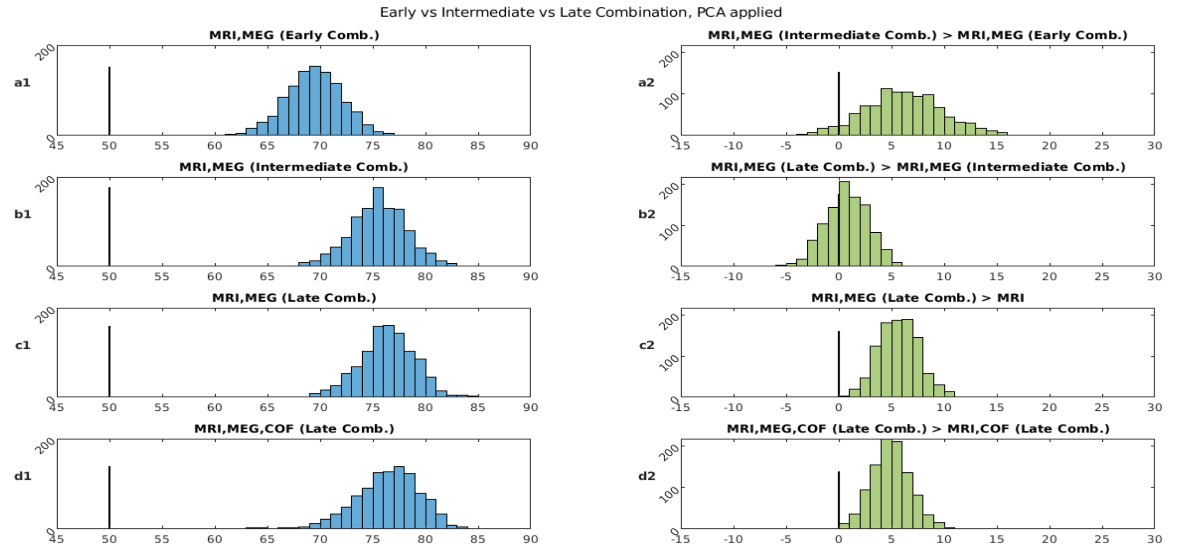


Supplementary Figure 6 Left column: Classification accuracies (chance = 50%) from 1000 random permutations using MEG, MRI with early, intermediate and late combinations (see methods), after applying PCA to the number of components within each modality needed to explain 95% of the variance across participants. Right column: Corresponding differences in classification performance for each permutation when comparing various combinations approach (where 0 = means no
